# Racial differences in white matter hyperintensity burden in aging, MCI, and AD

**DOI:** 10.1101/2022.04.11.22273726

**Authors:** Cassandra Morrison, Mahsa Dadar, Ana L. Manera, D. Louis Collins, Alzheimer’s Disease Neuroimaging Initiative

## Abstract

White matter hyperintensities may be one of the earliest pathological changes in aging and may potentially accelerate cognitive decline. Whether race influences WMH burden has been conflicting. The goal of this study was to examine if race differences exist in WMH burden and whether these differences are influenced by vascular factors [i.e., diabetes, hypertension, body mass index (BMI)]. Participants from the Alzheimer’s Disease Neuroimaging Initiative were included if they had a baseline MRI, diagnosis, and WMH measurements. Ninety-one Black and 1937 White individuals were included. Using bootstrap re-sampling, 91 Whites were randomly sampled and matched to Black participants based on age, sex, education, and diagnosis 1000 times. Linear regression models examined the influence of race on baseline WMHs with and without vascular factors: *WMH ∼ Race + Age + Sex + Education* + *BMI + Hypertension + Diabetes* and *WMH ∼ Race + Age + Sex + Education*. The 95% confidence limits of the t-statistics distributions for the 1000 samples were examined to determine statistical significance. All vascular risk factors had significantly higher prevalence in Black than White individuals. When not including vascular risk factors, Black individuals had greater WMH volume overall as well as in frontal and parietal regions, compared to White individuals. After controlling for vascular risk factors, no WMH group differences remained significant. These findings suggest that vascular risk factors are a major contributor to racial group differences observed in WMHs.

## 1 Introduction

Cerebrovascular disease (CVD) is a known risk factor for cognitive decline and dementia and may result in pathological brain changes [1]. These pathological brain changes are often characterized by *in vivo* MRI measures of white matter hyperintensities (WMH). Although WMHs are observed in cognitively healthy older adults, increased WMH load is associated with cognitive decline in normal aging, mild cognitive impairment (MCI), and dementia [2–5]. WMH burden may be of the earliest pathological changes in aging [6] and may lower the threshold for pathological aging [7] and diagnosis of dementia [8]. Vascular risk factors such as hypertension, diabetes, and body mass index (BMI) are implicated in increased WMH load, and are also independent risk factors of dementia and cognitive decline [8–10].

When examining WMH in a racially diverse cohort, results have been conflicting as to whether racial differences in WMH burden exist. For example, some studies report that there are no significant differences in WMH burden because of racial group (when comparing African American/Black individuals [hereafter referred to as Black(s) to European American/Caucasian/White individuals [hereafter referred to as White(s)]) [11,12]. Conversely, other studies have reported increased WMH burden in Blacks compared to Whites [13,14].

Further, higher deep brain WMH volume has also been observed in Blacks compared to Whites, particularly in those at risk for coronary artery disease [15]. The observed group differences may be associated with the strong relationship between WMHs and vascular burden [16], and between racially diverse samples exhibiting higher vascular burden compared to Whites. For example, several studies have shown that Blacks have a higher cerebrovascular disease burden compared to Whites [4,14,17]. The higher proportion of vascular risk factors in this population could be a contributing factor explicating why Black individuals are approximately twice as likely to develop dementia compared to White individuals [18–20].

While some of the aforementioned studies examine vascular risk factors in WMH, the results are still conflicting and require more research to understand whether vascular risk factors explain race differences in WMH burden. Some studies have reported racial differences in aging research, including healthy aging and dementia. Nevertheless, most dementia and aging research have been disproportionately conducted on non-Hispanic Whites [18]. This lack of diversity in research introduces challenges for both clinical and research settings by reducing the generalizability of findings in non-White populations. To understand factors important to cognitive decline and dementia, more studies are needed on racially diverse cohorts. Therefore, this study was designed to examine whether race influences overall and regional WMH burden in a sample of Black and White older adults. The analyses were designed to be conducted with and without controlling for vascular factors [i.e., diabetes, hypertension, body mass index (BMI)]. This design was completed to determine whether there are racial group differences in WMH load and whether these differences are explained by vascular factors. These findings will help improve the current understanding of how vascular factors contribute to WMH in a racially diverse sample.

## 2 Methods

### 2.1 Alzheimer’s Disease Neuroimaging Initiative

Data used in the preparation of this article were obtained from the Alzheimer’s Disease Neuroimaging Initiative (ADNI) database (adni.loni.usc.edu). The ADNI was launched in 2003 as a public-private partnership, led by Principal Investigator Michael W. Weiner, MD. The primary goal of ADNI has been to test whether serial magnetic resonance imaging (MRI), positron emission tomography (PET), other biological markers, and clinical and neuropsychological assessment can be combined to measure the progression of mild cognitive impairment (MCI) and early Alzheimer’s disease (AD). The study received ethical approval from the review boards of all participating institutions. Written informed consent was obtained from participants or their study partner. Participants were selected from all ADNI cohorts (ADNI-1, ADNI-2, ADNI-GO, and ADNI-3).

### 2.2 Participants

ADNI participant inclusion and exclusion criteria is available at www.adni-info.org. All participants were between the ages of 55 and 90 at the time of recruitment, exhibiting no evidence of depression. Healthy normal controls had no evidence of memory decline, as measured by the Wechsler Memory Scale and no evidence of impaired global cognition as measured by the Mini Mental Status Examination (MMSE) or Clinical Dementia Rating (CDR). MCI participants scored between 24 and 30 on the MMSE, 0.5 on the CDR, and abnormal scores on the Wechsler Memory Scale. Dementia was defined as participants who had abnormal memory function on the Wechsler Memory Scale, an MMSE score between 20 and 26 and a CDR of 0.5 or 1.0.

Participants were selected from the ADNI dataset for this study if they identified their race as “Black” or “White” and had completed baseline MRI scans. There were 140 Black individuals, of which 91 had baseline MRIs available. Out of the 2121 White individuals, 1937 had baseline MRIs available.

### 2.3 Structural MRI acquisition and processing

Baseline scans were downloaded from the ADNI public website. For the detailed MRI acquisition protocol and imaging parameters see http://adni.loni.usc.edu/methods/mri-tool/mri-analysis/.

T1w scans for each participant were pre-processed through our standard pipeline including noise reduction [21], intensity inhomogeneity correction (Sled, Zijdenbos, & Evans, 1998), and intensity normalization into range [0-100]. The pre-processed images were then linearly (9 parameters: 3 translation, 3 rotation, and 3 scaling) [22] registered to the MNI-ICBM152-2009c average [23]. The quality of the linear registrations was visually verified by an experienced rater (author M.D.), blinded to group. Only seven scans did not pass this quality control step and were discarded.

### 2.4 WMH measurements

A previously validated WMH segmentation technique was employed to generate participant WMH volume measurements. This segmentation technique has been validated in multi-center studies such as the Parkinson’s Progression Markers Initiative [24] and National Alzheimer’s Coordinating Center [25] Importantly, this technique has also been validated in ADNI [3] where a library of manual segmentations based on 50 ADNI participants (independent of those studied here) was created. WMHs were automatically segmented at baseline using the T1w contrasts, along with a set of location and intensity features obtained from a library of manually segmented scans in combination with a random forest classifier to detect the WMHs in new images [26,27]. The volumes of the WMHs for frontal, parietal, temporal, and occipital lobes as well as the entire brain were calculated based on Hammers atlas [26,28]. WMH load was defined as the volume of all voxels as WMH in the standard space (in mm^3^) and are thus normalized for head size. WMH volumes were log-transformed to achieve normal distribution.

### 2.5 Vascular Risk Factors

Vascular risk factors were included using information also downloaded from the ADNI public website. To calculate BMI for each person, height and weight information provided by ADNI for the matching visit to the MRI scan were used. Hypertension was defined by ADNI with each person being assigned a ‘0’ for no hypertension and a ‘1’ for hypertension. Missing documentation on diabetic status was completed using medication information. Medication lists were downloaded from ADNI, an experienced medical professional (author A.M.) identified all medications prescribed to manage diabetes. This list was then used as a proxy to determine which participants had diabetes.

### 2.6 Data availability statement

The data used for this analysis are available on request from the ADNI database (ida.loni.usc.edu).

### 2.7 Statistical Analysis

Analyses were performed using MATLAB R2019b. Linear regression models were conducted to examine whether Race would influence WMH burden. WMHs were examined in both a regional approach (frontal, temporal, parietal, occipital, left and right sides averaged together) for the entire brain. Analyses were completed separately for WMHs for each of the four regions and for the average of all regions for the whole brain.

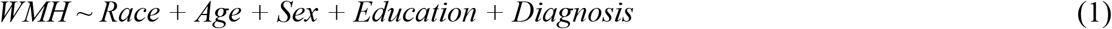

The categorical variable of interest was Race, indicated by either Black or White status based on participant self-identification. The model also included age, sex, years of education and diagnosis (categorical variable contrasting MCI and AD against the controls) as covariates. The analyses were also completed controlling for the vascular risk factors of diabetes, hypertension, and BMI.

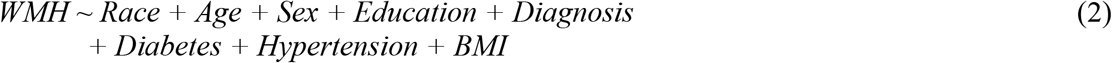

Again, the categorical variable of interest was Race, indicated by either Black or White status based on participant self-identification. The model also included age, sex, education, diagnosis, diabetes (categorical variable), hypertension (categorical variable), and BMI (continuous variable) as covariates.

To deal with the imbalance of data between groups, we opted to apply a bootstrapping procedure. This procedure subsamples the larger dataset multiple times and estimates the indirect effect in each resampled set [29]. We resampled 91 White participants from the original dataset of 1937 with replacement to get 1000 new datasets with the same size as the Black participant dataset. White participants were selected to match Black participants in sex, education, diagnosis, and age (age and education difference must have been less than 1 year). These analyses were completed with the 1000 bootstrapped samples. The 95% confidence limits of the t-statistics distributions for the 1000 samples were examined to determine whether the differences were statistically significant. To correct for multiple comparisons (N=10), we also examined the 99.5% confidence limits of t-statistics distributions for the 1000 samples.

To examine whether vascular risk factors differed between the groups, independent samples t-test and chi-square tests were completed. T-tests were completed to compare mean BMI, systolic blood pressure, and diastolic blood pressure between the groups. To examine differences in the number of people with diabetes and hypertension in each group, two chi-square tests were completed. A chi-square was also completed to examine Hachinski scores between the two groups.

## 3 Results

### 3.1 Descriptive Information of Vascular Factors

Table 1 presents the demographic and descriptive information for vascular factors for the two groups. White participants had higher education (*t*= 2.76, *p*=.006) and a higher ratio of males (*x*^*2*^=15.13, *p*=.003) than Black participants. Blacks had higher BMIs (*t*=3.21, *p*=.002), systolic blood pressure (*t*=2.34, *p*=.02), and diastolic blood pressure (*t*=2.19, *p*=.03) than Whites. The chi-square analyses found racial group differences for diabetes (*x*^*2*^=5.84, *p*=.016) and hypertension (*x*^*2*^=37.04, *p*<.001), with Blacks showing higher rates of both conditions than Whites. A chi-square analysis also revealed race differences for the Hachinski score (*x*^*2*^=38.00, *p*<.001) and diagnostic status (*x*^*2*^=8.75, *p=*.01). The significant effect in diagnostic status was because of the differences in the percentage of cognitively healthy older adults and those with MCI between the groups. There was a higher percentage of cognitive healthy older adult participants in the Black participant sample and a higher percentage of MCI participants in the White participant sample. Descriptive information for fasting glucose and cholesterol levels are also provided in Table 1. However, most participants were missing these values and therefore these vascular factors were not included in the regression models.

**Table 1:**
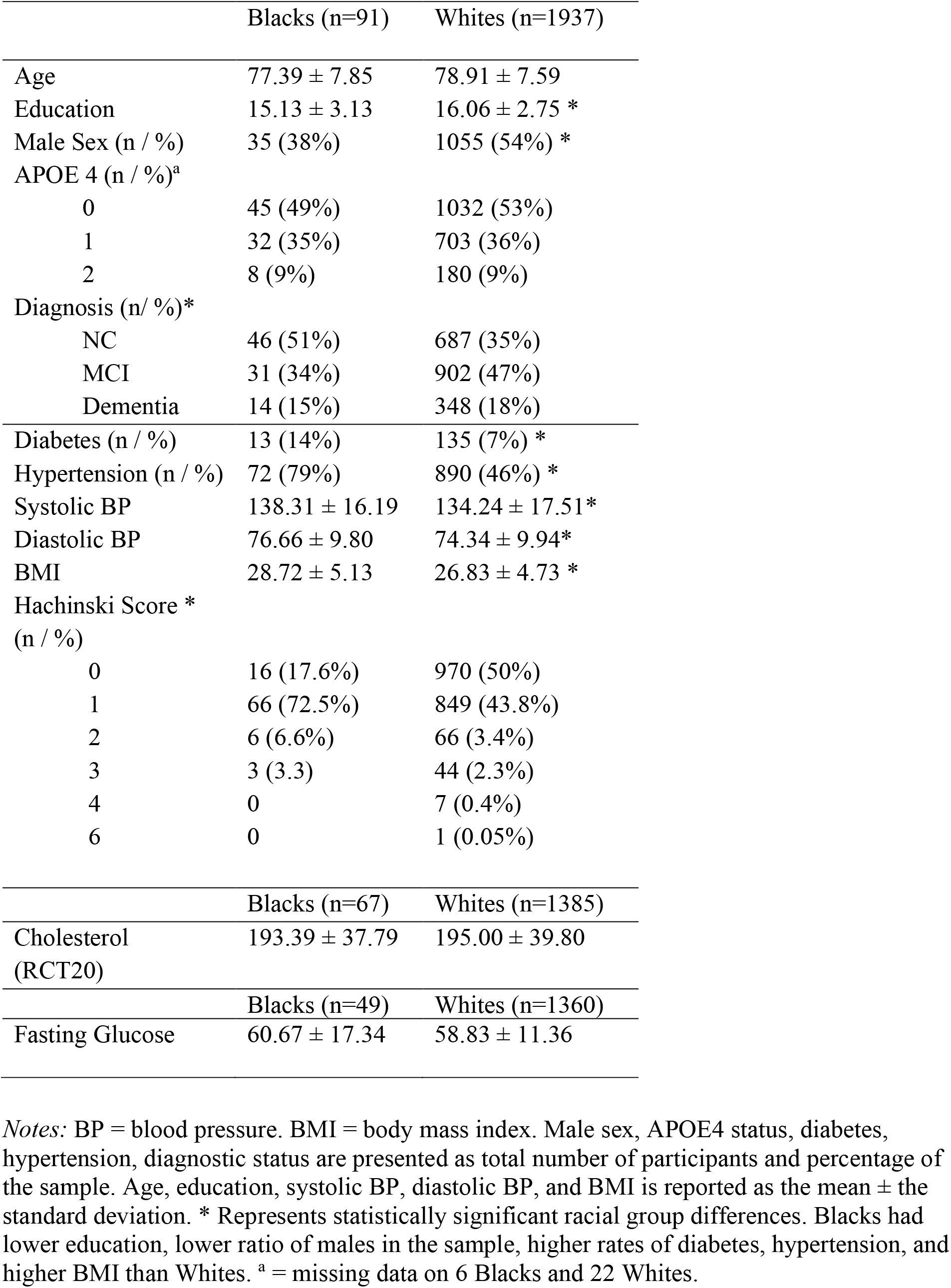
Descriptive information regarding vascular risk factors

|  | Blacks (n=91) | Whites (n=1937) |
| --- | --- | --- |
| Age | 77.39 ± 7.85 | 78.91 ± 7.59 |
| Education | 15.13 ± 3.13 | 16.06 ± 2.75 * |
| Male Sex (n / %) | 35 (38%) | 1055 (54%) * |
| APOE 4 (n / %)<sup>a</sup> |  |  |
| 0 | 45 (49%) | 1032 (53%) |
| 1 | 32 (35%) | 703 (36%) |
| 2 | 8 (9%) | 180 (9%) |
| Diagnosis (n/ %)* |  |  |
| NC | 46 (51%) | 687 (35%) |
| MCI | 31 (34%) | 902 (47%) |
| Dementia | 14 (15%) | 348 (18%) |
| Diabetes (n / %) | 13 (14%) | 135 (7%) * |
| Hypertension (n / %) | 72 (79%) | 890 (46%) * |
| Systolic BP | 138.31 ± 16.19 | 134.24 ± 17.51* |
| Diastolic BP | 76.66 ± 9.80 | 74.34 ± 9.94* |
| BMI | 28.72 ± 5.13 | 26.83 ± 4.73 * |
| Hachinski Score * |  |  |
| (n / %) |  |  |
| 0 | 16 (17.6%) | 970 (50%) |
| 1 | 66 (72.5%) | 849 (43.8%) |
| 2 | 6 (6.6%) | 66 (3.4%) |
| 3 | 3 (3.3) | 44 (2.3%) |
| 4 | 0 | 7 (0.4%) |
| 6 | 0 | 1 (0.05%) |
|  | Blacks (n=67) | Whites (n=1385) |
| Cholesterol (RCT20) | 193.39 ± 37.79 | 195.00 ± 39.80 |
|  | Blacks (n=49) | Whites (n=1360) |
| Fasting Glucose | 60.67 ± 17.34 | 58.83 ± 11.36 |

*Notes:* BP = blood pressure. BMI = body mass index. Male sex, APOE4 status, diabetes, hypertension, diagnostic status are presented as total number of participants and percentage of the sample. Age, education, systolic BP, diastolic BP, and BMI is reported as the mean ± the standard deviation. \* Represents statistically significant racial group differences. Blacks had lower education, lower ratio of males in the sample, higher rates of diabetes, hypertension, and higher BMI than Whites. <sup>a</sup> = missing data on 6 Blacks and 22 Whites.

### 3.2 WMH not controlling for vascular factors

Table 2 presents the median and lower and upper t-statistic confidence intervals obtained from the 1000 samples. T-statistics confidence intervals that do not include “0” indicate that WMH group differences due to race are statistically significant. Figure 1 shows the t-statistic and p-value for total WMH load both not controlling and controlling for vascular risk factors.

**Table 2:** Confidence intervals of the t-statistic for the 1000 iterations of Model 1 (i.e., not controlling for vascular risk factors).

|  | Median<br>Effect Size | Lower<br>95% CI | Upper<br>95% CI | Lower<br>99.5% CI | Upper<br>99.5% CI |
| --- | --- | --- | --- | --- | --- |
| Total WMH | -1.84 | -2.98 | -0.73* | -3.43 | -0.74* |
| Frontal WMH | -1.65 | -2.74 | -0.63* | -3.47 | -0.13* |
| Temporal WMH | 0.57 | -0.59 | 1.69 | -1.17 | 1.95 |
| Parietal WMH | -2.69 | -3.90 | -1.52* | -4.31 | -1.45* |
| Occipital WMH | -0.85 | -1.81 | 0.06 | -2.09 | 0.36 |

Notes: CI = confidence interval. WMH = white matter hyperintensity. \* = t-statistic is significant for group differences. n.s. = t-statistic is not significant for group differences.

**Figure 1:**
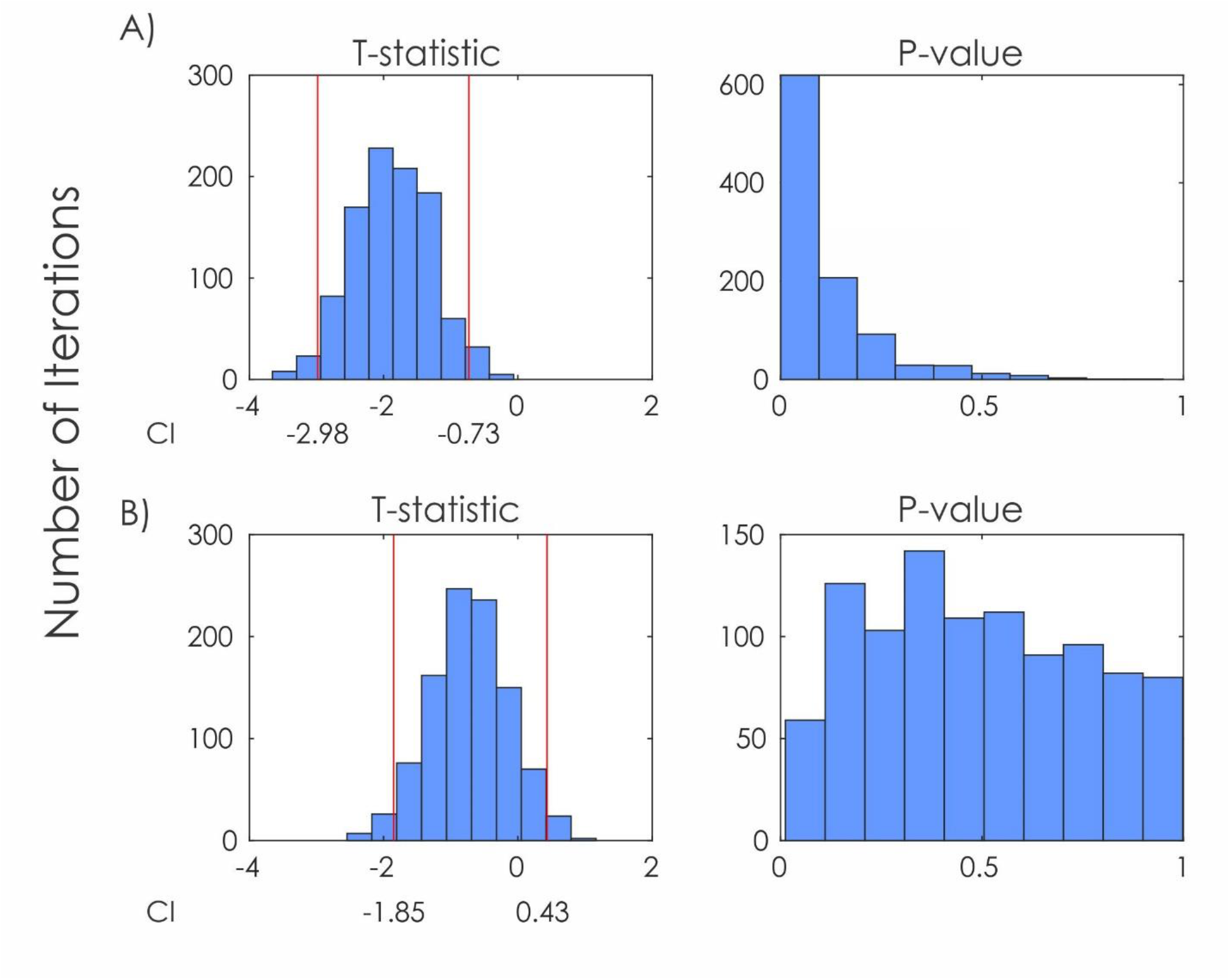
Image showing the t-statistic and p-value for each of the 1000 linear regressions examining effect of race on total WMH load. *Notes:* A) T-statistic and p-value for total WMH load controlling for age, sex, education, and diagnostic status. B) T-statistic and p-value for total WMH load controlling for age, sex, education, diagnostic status, BMI, hypertension, diabetes. CI = confidence intervals. Red lines represent the 95% CIs.

For total WMH, the lower and upper limits of the t-statistic CIs were -2.98 and -0.73, respectively. This finding indicates that total WMH burden was influenced by race. Black individuals had higher total WMH load compared to White individuals when controlling for age, sex, education, and diagnostic status. The frontal WMH CIs were -2.74 and -0.63, indicating significance and higher WMH burden in Black than White individuals. Parietal WMH load was also influenced by race (CIs = -3.90 and -1.52), with Black individuals exhibiting higher WMH load compared to White individuals. Neither temporal WMH (CIs = -0.59 and 1.69) or occipital WMH load (CIs = -1.81 and 0.06) significantly differed as a function of race. When examining the 99.5% confidence intervals (i.e., correcting for multiple comparisons) all results remained the same with total, frontal, and parietal WMH load remaining statistically different between the groups.

### 3.3 WMH controlling for vascular factors

Table 3 presents the median and lower and upper t-statistic confidence intervals obtained from the 1000 samples. T-statistics confidence intervals that do not include “0” indicate that WMH group differences due to race are statistically significant.

**Table 3:** Confidence intervals of the t-statistic for the 1000 iterations of Model 2 (i.e., controlling for vascular risk factors).

|  | Median<br>Effect Size | Lower<br>95% CI | Upper<br>95% CI | Lower<br>99.5% CI | Upper<br>99.5% CI |
| --- | --- | --- | --- | --- | --- |
| Total WMH | -0.70 | -1.85 | 0.43 | -2.33 | 0.80 |
| Frontal WMH | -0.63 | -1.81 | 0.48 | -2.42 | 1.00 |
| Temporal WMH | 0.91 | -0.29 | 2.22 | -0.57 | 2.54 |
| Parietal WMH | -1.62 | -2.83 | -0.43* | -3.46 | 0.17 |
| Occipital WMH | 0.27 | -0.69 | 1.22 | 1.17 | 1.46 |

*Notes:* CI = confidence interval. WMH = white matter hyperintensity. \* = t-statistic is significant for group differences. n.s. = t-statistic is not significant for group differences.

When controlling for vascular risk factors, only the racial group differences in WMH load remained significant in the parietal region (CIs = -2.83 and -0.43). That is, Black individuals had higher parietal WMH load compared to White individuals. This finding indicates that age, sex, education, and diagnosis do not account for differences in WMH between groups but that vascular risk factors drive most of the WMH group differences. Total (CIs = -1.85 and 0.43), frontal (CIs = -1.81 and 0.48), temporal (CIs = -0.29 and 2.22), and occipital WMH (CIs = -0.69 and 1.22), did not differ as a function of race. When examining the 99.5% confidence intervals (i.e., correcting for multiple comparisons) the parietal region was no longer statistically significant between the groups (CIs = -3.46 and 0.17).

## 4 Discussion

White matter hyperintensities (WMH) are known to develop with increased age and are strongly associated with cognitive decline and dementia [3]. These WMHs may occur as a result of cerebrovascular risk factors such as hypertension, diabetes, and increased BMI. Several studies have observed that Black individuals have increased cerebrovascular disease compared to White individuals [14]. As a result, one could predict that Black individuals would have a higher WMH load compared to Whites. However, whether racial differences exist in WMH burden has been conflicting. This study was designed to address limitations in current research by examining whether racial differences exist in WMH burden and whether they are influenced by vascular risk factors. The current findings suggest that Black individuals exhibit higher frontal, parietal, and total WMH load compared to Whites when vascular risk factors are not controlled for. On the other hand, when BMI, diabetes, and hypertension were included in the model, and correction for multiple comparisons was completed, significant group differences in total, frontal, and parietal WMH were no longer significant. The findings of the current study highlight important racial differences in aging research that have both research and clinical applications.

From a research perspective, these findings show that racial differences in aging, cognitive decline, and dementia may be accounted for by other underlying factors. In this study, racial group differences in WMH load were largely associated with the vascular risk factors examined. This finding is consistent with other studies that observed vascular disease is associated with higher WMH burden in Blacks, [11] and when controlling for vascular risk factors no racial group differences in WMH load are observed [16]. More research on racially/ethnically diverse populations is needed to fully understand neurodegeneration changes in aging and cognitive decline. Increasing research in multi-racial and multi-ethnic samples will make health research more generalizable and improve a doctors’ ability to treat and provide care to diverse populations.

From a clinical standpoint, previous research has shown that some ethnic groups report discrimination which leads to delays in medical testing, treatment, and receiving prescription medications [30] suggesting that there are racial and ethnic access disparities to health care [31]. Their conditions are thus more likely to be left untreated or undermanaged. Taken together with our current findings of higher WMH load in Blacks (than Whites), vascular risk factors may be undermanaged in this group leading to greater WMH load relative to White individuals. WMH may lower ones’ threshold for dementia and cognitive decline [7,8]. Thus, the higher WMH burden in Black compared to White individuals, may contribute to Blacks exhibiting an increased risk for and incidence of dementia compared to Whites [32]. Improved treatment and management of vascular risk factors in Black populations may reduce WMH burden and subsequent cognitive decline and dementia and should be studied in the future.

There are a few limitations in the current study that should be examined in future research. Other risk factors for cerebrovascular disease such as waist circumference, cholesterol, and fasting glucose were not included in the current study. ADNI did not conduct participant waist circumference measurements, while the fasting glucose and cholesterol participant data were missing for many participants limiting its applicability in the current sample. Future research should also examine healthcare and environmental exposure differences (e.g., social-economic status), which has been observed to be an important factor in racial differences in both vascular risk factors and WMH development [13]. Finally, the ADNI sample is comprised of well-educated participants which may limit the generalizability of the current findings to the general population.

## Conclusion

This study was novel in that it examined the influence of racial differences on WMHs while both controlling for and not controlling for vascular risk factors in the same sample of participants. We revealed that Blacks have more WMHs than Whites, which is influenced by them having increased BMI, more diabetes, and more hypertension than Whites. These findings suggest that racial differences in WMH burden are largely influenced by underlying vascular risk factors. Future research needs to determine the longitudinal implications of increased vascular risk factors in Blacks on WMH and both cognitive change and diagnostic status change over time.

## Data Availability

ALL data produced in this manuscript are available online at http://www.adni-info.org/

## Statements and Declarations

## Acknowledgments

Data collection and sharing for this project was funded by the Alzheimer’s Disease Neuroimaging Initiative (ADNI) (National Institutes of Health Grant U01 AG024904) and DOD ADNI (Department of Defense award number W81XWH-12-2-0012). ADNI is funded by the National Institute on Aging, the National Institute of Biomedical Imaging and Bioengineering, and through generous contributions from the following: AbbVie, Alzheimer’s Association; Alzheimer’s Drug Discovery Foundation; Araclon Biotech; BioClinica, Inc.; Biogen; Bristol-Myers Squibb Company; CereSpir, Inc.; Cogstate; Eisai Inc.; Elan Pharmaceuticals, Inc.; Eli Lilly and Company; EuroImmun; F. Hoffmann-La Roche Ltd and its affiliated company Genentech, Inc.; Fujirebio; GE Healthcare; IXICO Ltd.; Janssen Alzheimer Immunotherapy Research & Development, LLC.; Johnson & Johnson Pharmaceutical Research & Development LLC.; Lumosity; Lundbeck; Merck & Co., Inc.; Meso Scale Diagnostics, LLC.; NeuroRx Research; Neurotrack Technologies; Novartis Pharmaceuticals Corporation; Pfizer Inc.; Piramal Imaging; Servier; Takeda Pharmaceutical Company; and Transition Therapeutics. The Canadian Institutes of Health Research is providing funds to support ADNI clinical sites in Canada. Private sector contributions are facilitated by the Foundation for the National Institutes of Health (www.fnih.org). The grantee organization is the Northern California Institute for Research and Education, and the study is coordinated by the Alzheimer’s Therapeutic Research Institute at the University of Southern California. ADNI data are disseminated by the Laboratory for Neuro Imaging at the University of Southern California.

